# Potential Factors for Prediction of Disease Severity of COVID-19 Patients

**DOI:** 10.1101/2020.03.20.20039818

**Authors:** Huizheng Zhang, Xiaoying Wang, Zongqiang Fu, Ming Luo, Zhen Zhang, Ke Zhang, Ying He, Dongyong Wan, Liwen Zhang, Jing Wang, Xiaofeng Yan, Mei Han, Yaokai Chen

**Author notes:** Corresponding author: Prof. Dr. Yaokai Chen, Central Laboratory, Chongqing Public Health Medical Center, Chongqing 400036, China, Prof. Dr. Mei Han, Central Laboratory, Chongqing Public Health Medical Center, Chongqing 400036, China. These authors contribute equally to this work (Huizheng Zhang, Xiaoying Wang, Zongqiang Fu, Ming Luo, Mei Han, Yaokai Chen).

## Abstract

**Objective:** Coronavirus disease 2019 (COVID-19) is an escalating global epidemic caused by SARS-CoV-2, with a high mortality in critical patients. Effective indicators for predicting disease severity in SARS-CoV-2 infected patients are urgently needed.

**Methods:** In this study, 43 COVID-19 patients admitted in Chongqing Public Health Medical Center were involved. Demographic data, clinical features, and laboratory examinations were obtained through electronic medical records. Peripheral blood specimens were collected from COVID-19 patients and examined for lymphocyte subsets and cytokine profiles by flow cytometry. Potential contributing factors for prediction of disease severity were further analyzed.

**Results:** A total of 43 COVID-19 patients were included in this study, including 29 mild patients and 14 sever patients. Severe patients were significantly older (61.9±9.4 vs 44.4±15.9) and had higher incidence in co-infection with bacteria compared to mild group (85.7%vs27.6%). Significantly more severe patients had the clinical symptoms of anhelation (78.6%) and asthma (71.4%). For laboratory examination, 57.1% severe cases showed significant reduction in lymphocyte count. The levels of Interluekin-6 (IL6), IL10, erythrocyte sedimentation rate (ESR) and D-Dimer (D-D) were significantly higher in severe patients than mild patients, while the level of albumin (ALB) was remarkably lower in severe patients. Further analysis demonstrated that ESR, D-D, age, ALB and IL6 were the major contributing factors for distinguishing severe patients from mild patients. Moreover, ESR was identified as the most powerful factor to predict disease progression of COVID-19 patients.

**Conclusion:** Age and the levels of ESR, D-D, ALB and IL6 are closely related to the disease severity of COVID-19 patients. ESR can be used as a valuable indicator for distinguishing severe COVID-19 patients in early stage, so as to increase the survival of severe patients.

## Introduction

Coronaviruses possess a single strand, positive-sense RNA genome and belong to the family Coronaviridae. Most of the human coronaviruses are associated with mild clinical symptoms after infection^[1]^, except the two betacoronaviruses, severe acute respiratory syndrome coronavirus (SARS-CoV)^[2,3]^ and middle east respiratory syndrome coronavirus (MERS-CoV)^[4,5]^. Severe acute respiratory syndrome coronavirus 2 (SARS-CoV-2) is a newly recognised coronavirus emerged in Wuhan in December 2019^[6,7]^. It can cause a cluster of severe respiratory disease that has been recently named as Corona Virus Disease 2019 (COVID-19) by World Health Organization. As of March 18, 2020, the total number of confirmed COVID-19 patients are 80928 in the mainland of China and 3245 deaths were confirmed^[8]^.

Recent studies about the clinical characteristics of COVID-19 patients showed that the common clinical symptoms were fever, cough, fatigue and pneumonia for mild patients, while severe patients could develop to acute respiratory distress syndrome and even organ failure^[6,9,10]^. Patients with moderate symptoms, especially the elderly or the ones with comorbidity, can worsen and result in high mortality^[6,10]^. Several studies have reported the role of inflammation-related biomarkers during disease progression of COVID-19 patients. A study about 52 critically ill patients with SARS-CoV-2 pneumonia have demonstrated that lymphocytopenia reflects the severity of SARS-CoV-2 infection^[11]^. Consistent with this study, Li Tan et al. suggest that lymphocytopenia can be used as an useful prognostic factor for severe COVID-19 patients^[12]^. Recent studies have also demonstrated the decreases of lymphocytes and the increases of inflammatory cytokine levels in COVID-19 patients^[9,10]^ and further suggest neutrophil-to-CD8^+^ T cell ratio (N8R)^[13]^ and IL6 level^[14]^ as an indicator for early identification of severe COVID-19 patients. In line with above studies, Huang C et al. report that patients in intensive care unit (ICU) manifest higher expression of cytokines, including IL-2, IL-7, IL-10, GSCF and TNF, compared with non-ICU patients^[6]^. However, the correlation between the expression of clinical parameters and the disease severity is still largely unclear. In this study, we aimed to analyze the different characteristics of lymphocyte subsets and cytokine levels between mild and severe patients, and further screen suitable indicators for predicting disease severity, in order to provide guidance for subsequent clinical intervention.

## Method

### Study design and patients

For this retrospective study, 43 COVID-19 patients at Chongqing Public Health Medical Center from February 11, 2020 to February 28, 2020 were enrolled. Chongqing Public Health Medical Center is one of the four designated hospital in Chongqing for COVID-19 patients, mainly responsible for the treatment of COVID-19 patients from the main city area. All the involved COVID-19 patients were diagnosed according to the Guidelines of the Diagnosis and Treatment of Novel Coronavirus Pneumonia (version fifth) published by the National Health Commission of China and divided into mild and severe groups^[15]^. The mild patients should have all of the following conditions: fever or other respiratory symptoms, CT image with typical viral pneumonia and SARS-CoV-2 RNA positive by RT-PCR examination. For severe patients, at least one of the following conditions should be additionally met: 1) respiratory distress, RR ≥30 times/minute, 2) oxygen saturation ≤93% under the resting state, 3) oxygen partial pressure (PaO2)/oxygen concentration (FiO2) in arterial blood≤300mmHg. The study was approved by Chongqing Public Health Medical Center Ethics Committee and written informed consent was waived due to the anonymous analysis of the clinical data.

### Data collection

We obtained the clinical features of COVID-19 patients from the Center’s electronic medical records, including demographic information, clinical symptoms, laboratory analyses. Clinical laboratory investigation included whole blood count, serum biochemical test (including liver and renal function, lactate dehydrogenase, electrolytes and creatine kinase), coagulation test, C-reactive protein and procalcitonin. As for SARS-CoV-2 nucleic acid test, respiratory specimens, including throat swab specimens, bronchoalveolar lavage fluid or sputum were tested for SARS-CoV-2 RNA by using real time RT-PCR kits (Chongqing Zhongyuanhuiji Biotechnology Limited, Chongqing, China) recommended by the Chinese Center for Disease Control and Prevention (CDC). Bacterial, fungal and pathogenic microorganism examination was also routinely performed. All data were checked independently by two researchers to ascertain the accuracy.

### Lymphocyte subsets and cytokine profiles examination

Peripheral blood samples were collected from the involved COVID-19 patients for lymphocyte subsets and cytokines test. The lymphocyte test kit (Hangzhou CellGene Biotechnology Limited, Hangzhou, China) was used for lymphocyte subset analysis. Cytokines profiles (IL2, IL4, IL6, IL10, TNF-α, IFN-γ and IL17A) were detected with human Th1/2/17 cytokine kit (Jiangxi CellGene Biotechnology Limited, Jiangxi, China). All tests were performed according to the product manual.

### Statistical analysis

Categorical variables were expressed as the counts and percentages (%), and significance was detected by Fisher’s exact test in different categories. The quantized variables of parameters were described as mean and standard deviation if they were normally distributed, and the significance was tested by t-test; or expressed in median and quartile intervals if they were not normally distributed, and significance was tested by Wilcoxon rank-sum test. A two-sided p value less than 0.05 was considered statistically significant. Least absolute shrinkage and selection operator (Lasso) analysis was used to identify the major contributing factors among clinical parameters to differentiate mild and severe COIVD-19 patients. The predictive values of selected parameters for distinguishing COVID-19 patients with mild and severe symptoms were evaluated by receiver operating characteristic (ROC) and area under the ROC curve (AUC). Statistical analyses were done using the Stata software (version 15.0) and R package (version 3.5.3).

## Results

### Epidemiological and clinical characteristics of COVID-19 patients

A total of 43 COVID-19 patients were enrolled in this study, including 29 mild patients (67.4%) and 14 sever patients (32.6%) according to above mentioned conditions. The 43 COVID-19 patients consisted of 21 female patients (48.8%) and 22 male patients (51.2%). The medium ages of severe patients were significantly higher than that of mild patients (61.9±9.4 vs 44.4±15.9, p<0.001). About half of the patients (20, 46.5%) had exposure history, including have been to Wuhan (14, 32.6%) or had a contact history with the personnel returned from Wuhan (6, 13.9%). A total of 12 patients had underlying diseases, including hypertension (4, 9.3%), diabetes (6, 14.0%), tuberculosis (1, 2.3%) and rheumatic heart (1, 2.3%) and no significant differences were observed between mild and severe groups. Severe patients had much higher opportunity of bacterial co-infection than mild patients (85.7%vs27.6%, p<0.001). All the severe patients and 65.5% mild patients had fever, and significantly more severe patients had the temperature degree more than 39°C (42.9%vs6.9%, p=0.004). For the clinical symptoms, the severe patients had much higher frequencies of occurrence of chest distress (p=0.046), anhelation (p=0.008) and asthma (p=0.002) (Table 1).

**Table 1.** Demographic and clinical characteristics of COVID-19 patients

| Baseline variables | All patients<br>(N=43) | Mild patients<br>(N=29) | Severe patients<br>(N=14) | P-value |
| --- | --- | --- | --- | --- |
| **Characteristics** |  |  |  |  |
| Age (year), mean (SD) |  | 44.34±15.84 | 61.7±9.22 | <0.001 |
| Gender (%) |  |  |  | 0.20 |
| Women | 21 (48.8%) | 12 (41.4%) | 9 (64.3%) |  |
| Men | 22 (51.2%) | 17 (58.6%) | 5 (35.7%) |  |
| Returned from Wuhan | 14 (32.6%) | 10 (34.5%) | 4 (28.6%) | 1.00 |
| Contact with the<br>personnel returned from<br>Wuhan | 6 (14.0%) | 4 (14%) | 2 (14%) | 1.00 |
| **Underlying diseases**<br>**(%)** |  |  |  |  |
| Hypertension | 4 (9.3%) | 3 (10.3%) | 1 (7.1%) | 1.00 |
| Diabetes | 6 (14.0%) | 3 (10.3%) | 3 (21.4%) | 0.37 |
| Tuberculosis | 1 (2.3%) | 1 (3.4%) | 0 (0.0%) | 1.00 |
| Rheumatic heart | 1 (2.3%) | 1 (3.4%) | 0 (0.0%) | 1.00 |
| **Co-infection** |  |  |  |  |
| Bacterial | 20 (46.5%) | 8 (27.6%) | 12 (85.7%) | <0.001 |
| Fungal | 1 (2.3%) | 0 (0.0%) | 1 (7.1%) | 0.33 |
| **Highest**<br>**temperature, °C** |  |  |  |  |
| <37.3 | 10(23.2%) | 10(34.5%) | 0(0.0%) |  |
| 37.3-38.0 | 11(25.6%) | 7 (24.1%) | 4 (28.6%) | 0.118 |
| 38.1-39.0 | 14(32.6%) | 10 (34.5%) | 4 (28.6%) | 0.195 |
| >39.0 | 8(18.6%) | 2 (6.9%) | 6 (42.9%) | 0.004 |
| **Symptoms** |  |  |  |  |
| Chilly | 13 (30.2%) | 9 (31.0%) | 4 (28.6%) | 1.00 |
| Shiver | 3 (7.0%) | 3 (10.3%) | 0 (0.0%) | 0.54 |
| Fatigue | 17 (39.5%) | 13 (44.8%) | 4 (28.6%) | 0.34 |
| Cough | 39 (90.7%) | 26 (89.7%) | 13 (92.9%) | 1.00 |
| Phlegm | 36 (83.7%) | 24 (82.8%) | 12 (85.7%) | 1.00 |
| Pharyngeal itching | 5 (11.6%) | 5 (17.2%) | 0 (0.0%) | 0.16 |
| Pharyngalgia | 5 (11.6%) | 5 (17.2%) | 0 (0.0%) | 0.16 |
| Swirl | 12 (27.9%) | 7 (24.1%) | 5 (35.7%) | 0.48 |
| Headache | 8 (18.6%) | 6 (20.7%) | 2 (14.3%) | 1.00 |
| Nasal obstruction | 5 (11.6%) | 5 (17.2%) | 0 (0.0%) | 0.16 |
| Rhinorrhea | 8 (18.6%) | 7 (24.1%) | 1 (7.1%) | 0.24 |
| Chest distress | 15 (34.9%) | 7 (24.1%) | 8 (57.1%) | 0.046 |
| Pectoralgia | 6 (14.0%) | 5 (17.2%) | 1 (7.1%) | 0.65 |
| Anhelation | 20(46.5%) | 9 (31.0%) | 11 (78.6%) | 0.008 |
| Asthma | 16 (37.2%) | 6 (20.7%) | 10 (71.4%) | 0.002 |
| Myalgia | 6 (14.0%) | 6 (20.7%) | 0 (0.0%) | 0.15 |
| Abdominal pain | 3 (7.0%) | 3 (10.3%) | 0 (0.0%) | 0.54 |
| Diarrhea | 14 (32.6%) | 8 (27.6%) | 6 (42.9%) | 0.49 |
| Nausea | 4 (9.3%) | 4 (13.8%) | 0 (0.0%) | 0.29 |
| Emesis | 1 (2.3%) | 1 (3.4%) | 0 (0.0%) | 1.00 |

Note: Data are Mean (SD), Median (IQR), n (%). P values compare mild and severe patients using student t test, Fisher's exact test and Wilcoxon rank-sum test.

### Laboratory parameters of COVID-19 patients

For lymphocyte subsets detection, significantly decreases in CD3 (p=0.002), CD4 (p=0.002) and CD8 (p=0.009) T cell counts were observed in severe patients, while no significant difference in CD4^+^/CD8^+^ ratio between the two groups. For routine blood tests, severe patients showed significantly increased number (p=0.043) and percentage (p=0.001) of neutrophil and significantly decreased number and percentage of lymphocyte as well as eosinophil (lymphocyte: p=0.022 for number, p=0.005 for percentage; eosinophil: p=0.007 for number, p=0.006 for percentage). Lymphopenia (lymphocyte count <1.0×10^9^/L) was found in 8 (57.1%) severe patients and 10 (34.5%) mild patients. Furthermore, severe patients showed a significant reduction in hemoglobin (HGB, p=0.002), hematocrit (HCT, p=0.002) and the number of RBC (p=0.001) compared to mild patients. For coagulation function, level of activated partial thromboplastin time (APTT, p=0.043) in severe patients was significantly lower, while the levels of D-Dimer (D-D, p<0.001) and fibrinogen (FIB, p=0.024) were markedly greater in severe patients (Table 2).

**Table 2.** Analysis of laboratory examination at the admission of involved COVID-19 patients

| Baseline variables | Mild patients<br>(N=29) | Severe patients<br>(N=14) | P-value |
| --- | --- | --- | --- |
| **Lymphocyte subsets** |  |  |  |
| CD3 (cell/μL), mean (SD) | 828.69 ± 419.48 | 430.786 ± 209.71 | 0.002 |
| CD4 (%), mean (SD) | 55.76 ± 9.66 | 52 ± 10.59 | 0.250 |
| CD4 (cell/uL), mean (SD) | 456.38 ± 238.76 | 226.86 ± 132.64 | 0.002 |
| CD8 (%), mean (SD) | 40.27 ± 9.16 | 44.28 ± 10.75 | 0.210 |
| CD8 (cell/uL), mean (SD) | 339.45 ± 194.95 | 188.14 ± 97.09 | 0.009 |
| CD4/CD8, median (IQR) | 1.37 (1.08, 1.99) | 1.15 (0.76, 1.76) | 0.300 |
| **Infection-related biomarkers** |  |  |  |
| IFN-γ (pg/ml), median (IQR) | 1.44 (1.02, 1.89) | 2.05 (1.12, 2.65) | 0.130 |
| TNF-α (pg/ml), median (IQR) | 1 (0, 1.64) | 1.28 (0.95, 2.12) | 0.120 |
| IL-10 (pg/ml), median (IQR) | 2.35 (1.45, 3.87) | 5.78 (3.87, 7.36) | <0.001 |
| IL-6 (pg/ml), median (IQR) | 3.59 (1.38, 8.68) | 16.62 (2.39, 46.21) | 0.041 |
| IL-4 (pg/ml), median (IQR) | 0.9 (0.13, 1.26) | 0.99 (0.53, 1.61) | 0.41 |
| IL-2 (pg/ml), median (IQR) | 1 (0.9, 1.29) | 1.26 (0.85, 1.61) | 0.590 |
| IL-17A (pg/ml), median (IQR) | 10.03 (0, 19.51) | 17.74 (8.22, 25.34) | 0.150 |
| CRP (mg/L), median (IQR) | 5.6 (2.9, 21.95) | 41.66 (22.58, 61.33) | <0.001 |
| ESR (mm/h), median (IQR) | 19.93 ± 16.07 | 72.36 ± 28.10 | <0.001 |
| PCT (ng/ml), median (IQR) | 0.021 (0.02, 0.046) | 0.0325 (0.02, 0.075) | 0.430 |
| **Coagulation function** |  |  |  |
| PT (s), median (IQR) | 10.4 (10.1, 10.7) | 10.5 (10.3, 11) | 0.310 |
| INR, median (IQR) | 0.9 (.87, .92) | 0.91 (.89, .95) | 0.290 |
| APTT (s), mean (SD) | 26.93±2.49 | 25.13 ±2.97 | 0.043 |
| TT(s), mean (SD) | 16.89±1.46 | 16.56±0.75 | 0.430 |
| D-D (µg/L), median (IQR) | 0.34 (0.2, 0.51) | 2.22 (0.57, 6.84) | <0.001 |
| FIB(g/L), mean (SD) | 3.50±1.35 | 4.70 ±1.98 | 0.024 |
| **Blood routine** |  |  |  |
| WBC(×10<sup>9</sup>/L), median (IQR) | 5.13 (4.25, 6.26) | 5.91 (3.9, 10.08) | 0.200 |
| Neutrophil (×10<sup>9</sup>/L), median (IQR) | 2.86 (2, 3.5) | 4.77 (2.68, 7.33) | 0.043 |
| Neutrophil (%), mean (SD) | 59.16±14.52 | 74.71±12.50 | 0.001 |
| Lymphocyte (×10<sup>9</sup>/L) | 1.50±0.69 | 1.01±0.45 | 0.022 |
| Lymphocyte (%), mean (SD) | 30.16±13.56 | 17.83±11.21 | 0.005 |
| Monocyte (×10<sup>9</sup>/L), median (IQR) | 0.37 (0.32, 0.48) | 0.40 (0.29,0.56) | 0.900 |
| Monocyte (%), median (IQR) | 7.1 (6.2, 10.2) | 6.7 (3.8, 8.6) | 0.170 |
| Eosinophil (×10<sup>9</sup>/L), median (IQR) | 0.02 (0.01, 0.13) | 0 (0, 0.01) | 0.007 |
| Eosinophil (%), median (IQR) | 0.5 (0.1, 2.4) | 0.05 (0,0 .1) | 0.006 |
| RBC (×10<sup>12</sup>/L), mean (SD) | 4.45±0.41 | 3.96±0.46 | 0.001 |
| HGB (g/L), mean (SD) | 136.69 ±13.47 | 121.57±16.07 | 0.002 |
| PLT (×10<sup>9</sup>/L), mean (SD) | 170 (135, 232) | 186 (126, 270) | 0.560 |
| **Blood biochemistry** |  |  |  |
| PA (U/L), mean (SD) | 220.24±66.03 | 160.93±49.62 | 0.005 |
| AFU (U/L), mean (SD) | 24.88±7.17 | 29.66±9.93 | 0.079 |
| ALT (U/L), median (IQR) | 24 (16, 43) | 38 (20, 51) | 0.250 |
| AST (U/L), median (IQR) | 25 (19, 34) | 37 (26, 50) | 0.007 |
| AST/ALT, median (IQR) | 1 (0.7, 1.2) | 0.95 (0.8, 1.7) | 0.680 |
| ALP (U/L), median (IQR) | 51 (44, 64) | 60 (46, 74) | 0.320 |
| GGT (U/L), median (IQR) | 40 (18, 53) | 39.5 (21, 87) | 0.310 |
| LDH (U/L), median (IQR) | 211 (185, 236) | 311 (273, 420) | <0.001 |
| TP (g/L), mean (SD) | 68.28±7.31 | 67.18±6.88 | 0.640 |
| ALB (g/L), mean (SD) | 42.85±4.30 | 38.05±5.01 | 0.002 |
| GLO (g/L), median (IQR) | 24.5 (22, 27.7) | 29.1 (27.7, 30.3) | 0.006 |
| TBIL (µmol/L), mean (SD) | 14.90±7.60 | 15.4 ±4.67 | 0.830 |
| DBIL (µmol/L), mean (SD) | 4.97±2.70 | 5.17±1.58 | 0.800 |
| IDBIL(µmol/L), mean (SD) | 9.92±4.97 | 10.22±3.33 | 0.840 |
| TBA (µmol/L), median (IQR) | 2.8 (2.1, 6.1) | 6.7 (3.1, 9) | 0.110 |
| UREA (mmol/L), median (IQR) | 3.6 (3.16, 5.05) | 3.785 (3.29, 4.46) | 0.770 |
| CREA (µmol/L), mean (SD) | 68.91±17.58 | 66.6±17.05 | 0.690 |
| BMG (mg/L), median (IQR) | 2.34 (1.58, 2.91) | 2.815 (2.16, 3.66) | 0.062 |
| Cysc (mg/L), median (IQR) | 0.96 (0.9, 1.09) | 1.115 (0.97, 1.16) | 0.018 |
| eGFR, median (IQR) | 102 (95, 112) | 92 (79, 99) | 0.011 |
| CK (U/L), median (IQR) | 78 (52, 127) | 93.5 (51, 199) | 0.820 |
| CKMB ($\mu\text{g/L}$), median (IQR) | 9.2 (8.2, 11.1) | 10.1 (7.4, 12.6) | 0.740 |
| HBDH (U/L), median (IQR) | 157 (141, 177) | 240 (180, 314) | 0.002 |
| 5-NT (U/L), median (IQR) | 4.1 (2.8, 5) | 5.2 (4.3, 9) | 0.017 |
| TG (mmol/L), median (IQR) | 1.3 (1.02, 1.53) | 1.62 (1.39, 2.83) | 0.037 |
| CHOL (mmol/L), median (IQR) | 4.4 (3.69, 4.61) | 5.095 (4.04, 5.8) | 0.110 |
| HDL (mmol/L), mean (SD) | 1.08±0.24 | 1.10±0.20 | 0.790 |
| LDL (mmol/L), mean (SD) | 2.35±0.61 | 2.725±0.70 | 0.085 |
| K (mmol/L), median (IQR) | 4.09 (3.72, 4.45) | 3.685 (3.45, 3.92) | 0.035 |
| NA (mmol/L), median (IQR) | 138.1 (135.9, 139.9) | 138.1 (137, 141.4) | 0.780 |
| CL (mmol/L), median (IQR) | 103.5 (100.6, 106.2) | 100.55 (100, 105.1) | 0.320 |
| CA (mmol/L), median (IQR) | 2.25 (2.15, 2.3) | 2.185 (2.13, 2.27) | 0.360 |
| P (mmol/L), mean (SD) | 1.10±0.21 | 0.97±0.25 | 0.078 |
| MG (mmol/L), median (IQR) | 0.88 (0.84, 0.93) | 0.89 (0.79, 0.98) | 0.690 |
| CO2 (mmol/L), median (IQR) | 25 (23, 26) | 23.5 (23, 26) | 0.800 |
| Glu (mmol/L), median (IQR) | 6 (5.61, 6.32) | 6.76 (5.98, 8.02) | 0.007 |

Note: Data are Mean (SD) and Median (IQR). P values compare mild and severe patients using student t test, Fisher's exact test and Wilcoxon rank-sum test.

For the liver and kidney function, severe patients showed significantly higher expressions of aspartate aminotransferase (AST, p=0.007), lactate dehydrogenase, (LDH, p<0.001), globulin (GLO, p=0.006), cystatin c (cysc, p=0.018), hydroxybutyric dehydrogenase (HBDH, p=0.002), 5′-nucleotidase (5-NT, p=0.017), thyrogloblin (TG, p=0.037) and glucose (Glu, p=0.007), while showed significantly lower levels of albumin (ALB, p=0.002), albumin/globulin (A/G, p<0.001), uric acid (UA, p=0.002), glomerular filtration rate (eGFR, p=0.011) and K (p=0.035) compared with mild group (Table 2).

For the infection index, expression of C-reactive protein (CRP, p<0.001) and erythrocyte sedimentation rate (ESR, p<0.001) was significantly elevated in severe patients than mild patients. Further analysis of plasma cytokines demonstrated that the median concentrations of IFN-γ, TNF-α, IL10, IL6, IL4, IL2 and IL17A in severe group were higher than mild group. Levels of IL10 (p<0.001) and IL6 (p=0.041) increased significantly compared to mild patients. No significant differences in the levels of IFN-γ, TNF-α, IL4, IL2 and IL17A were observed between the two groups (Table 2).

Furthermore, neutrophil-to-CD8+ T cell ratio (N8R, p=0.002), neutrophil-to-lymphocyte ratio (NLR, p=0.006), the ratio of neutrophil ratio-to-lymphocyte ratio (NRLR, p=0.005) were significantly higher in severe patients than mild patients; while the CD3-WBC ratio (CD3W) was significantly decreased in severe patients (p<0.001) (data not show).

### Factors for the prediction of severe COVID-19 patients

We next identified the potential contributing factors to distinguish mild and severe patients by using the above mentioned parameters. 3-fold cross-validation via minimum criteria was firstly performed to define the tuning parameter in the LASSO model (Figure 1A). Further LASSO binary logistic regression analysis of 85 clinical related parameters indicated that age, IL6, ESR, ALB and D-D were closely related to disease severity of COVID-19 patients (Figure 1B). To assess the diagnostic value of the five selected factors, receiver operating characteristic (ROC) curve and area under ROC curve (AUC) were calculated by R package (Figure 2). The results indicated that ESR had a higher AUC (0.951) than D-D (0.892), age (0.819), ALB (0.760) and IL6 (0.695). Further analysis indicated that significantly differences in ROC values were observed between ESR and age (p=0.047), ESR and IL6 (p=0.009), ESR and ALB (p=0.021), except ESR and D-D (p=0.231).

**Figure 1.**
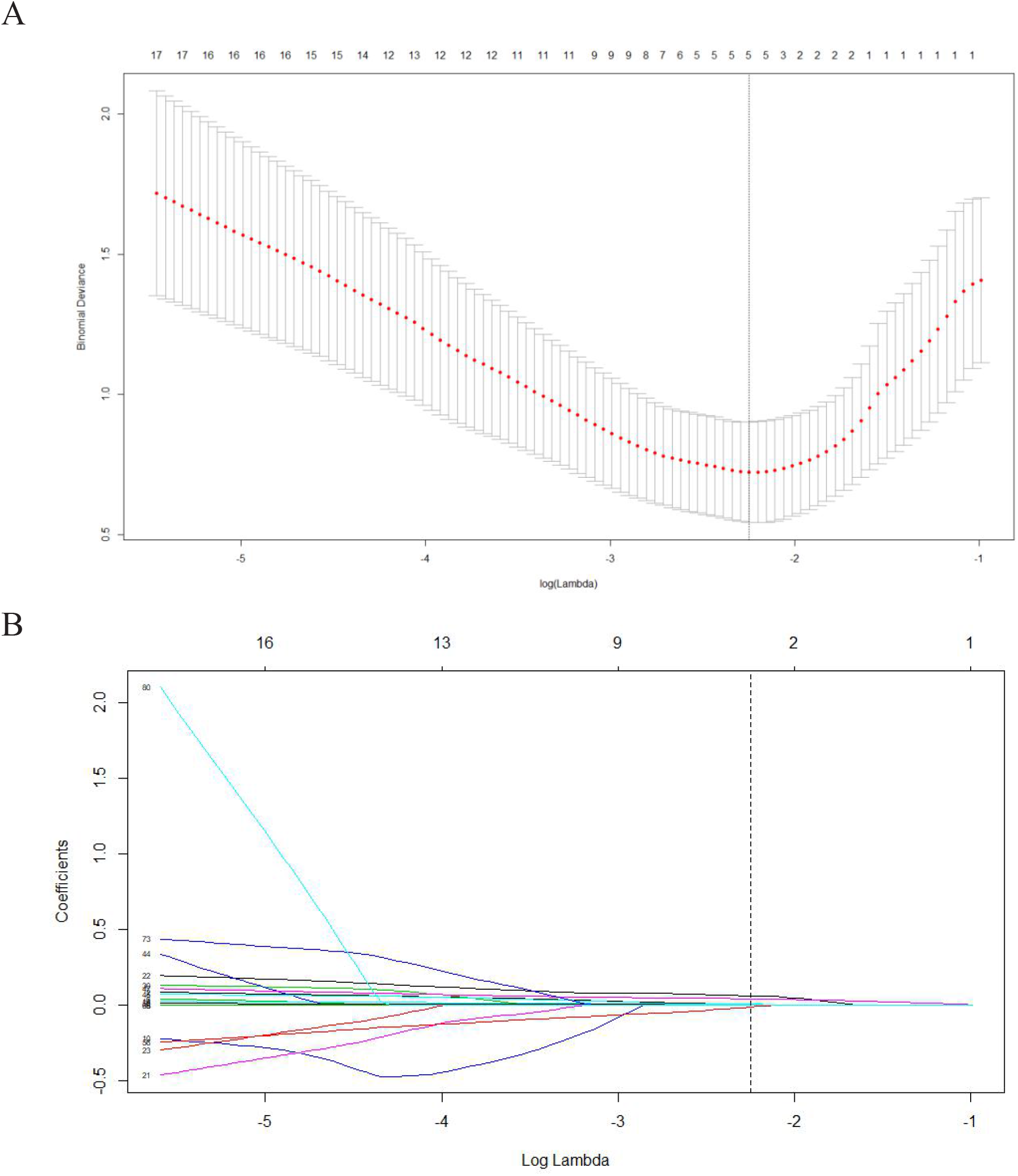
Selection of potential contributing factor using the least absolute shrinkage and selection operator (LASSO) binary logistic regression model. (A) lambda parameter selection in the LASSO model used 3-fold cross-validation via minimum criteria. Dotted vertical lines were drawn at the optimal values by using the minimum criteria. A lambda value of 0.108, where the dotted line is located, was chosen according to 3-fold cross-validation. (B) LASSO coefficient profiles of the 85 clinical parameters. Every line was the variation curve of the coefficient profile that changed with the lambda value. Vertical line was drawn at the value selected using 3-fold cross-validation, where optimal lambda resulted in 5 nonzero coeffificients, including erythrocyte sedimentation rate (ESR), D-Dimer, age, albumin (ALB) and interluekin-6 (IL6).

**Figure 2.**
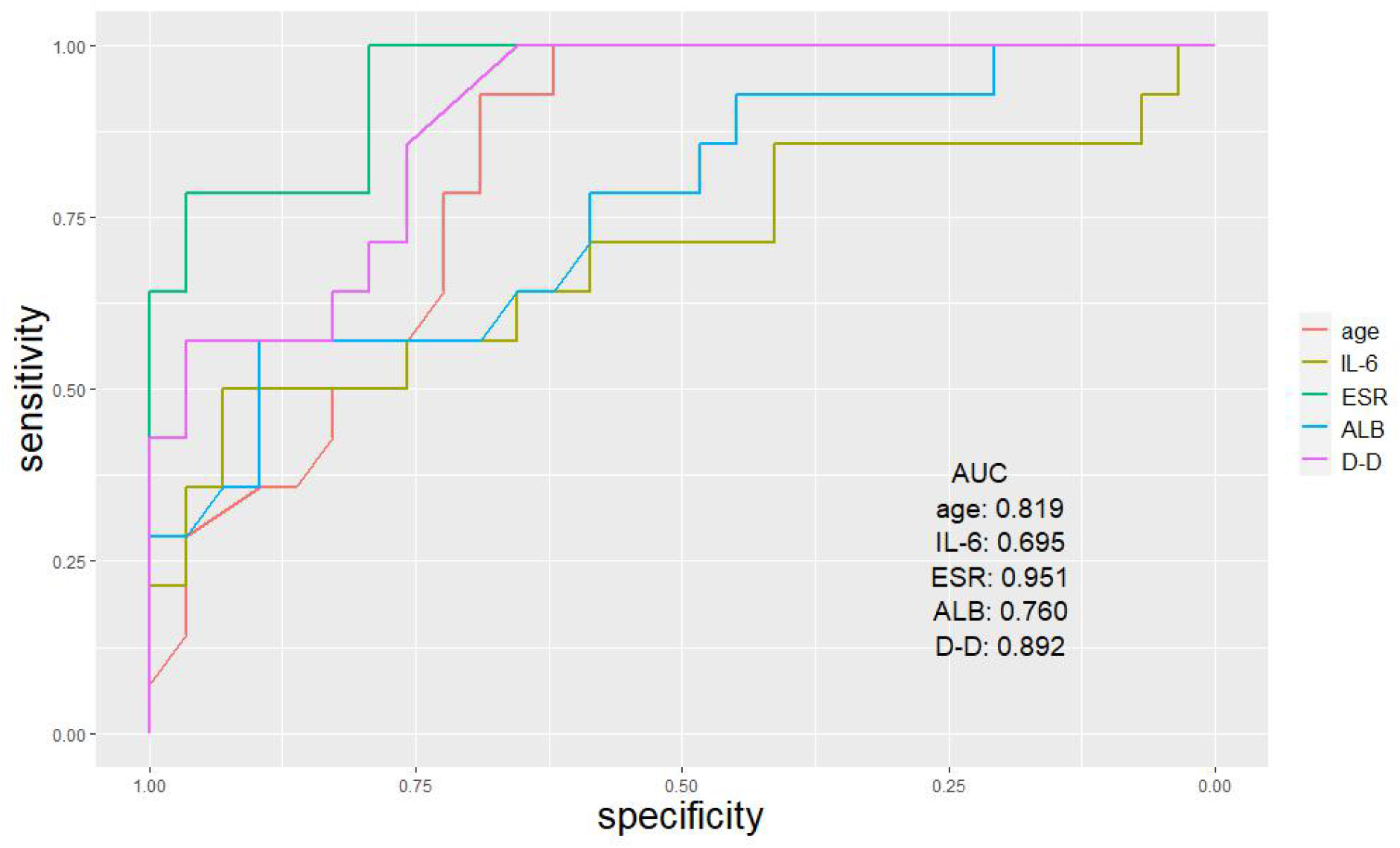
Assessment of the diagnostic value of selected contributing factors. ROC curve and AUC were calculated for the 5 selected parameters by using R package. The AUC values of the 5 parameters was 0.951 (ESR), 0.892 (D-Dimer), 0.819 (age), 0.760 (ALB) and 0.695 (IL6).

## Discussion

Recently, several studies focus on the immunologic characteristics and potential factors for monitoring disease severity in COVID-19 patients^[12,13,14,16, 17]^. However, knowledge about the role of clinical parameters involved in COVID-19 patients is still insufficient. In this study, we evaluated the clinical and immunological characteristics of 43 confirmed COVID-19 patients admitted at Chongqing Public Health Medical Center, Chongqing. And we further screened potential factors as effective indicators for disease progression, in order to provide appropriate treatment for severe patients early. Compared to mild patients, severe patients were older and had higher frequencies to be co-infected with bacteria, which is consistent with previous reports that critically ill patients were more likely to be older and had co-infection of bacteria^[9]^.

Furthermore, severe patients were more likely to have chest distress and respiratory symptoms, such as anhelation and asthma. The clinical symptoms of COVID-19 patients are similar to those reported by other studies^[6,9,10]^. In terms of laboratory finding, IL6, IL10, ESR, CRP, APTT, D-D, ALB, AST, LDH and FIB were significantly higher in severe cases than mild cases, which indicate a degree of cardiac, live, coagulation function abnormality and infection, so more attention should be paid for these patients to avoid delayed treatment.

In our study, severe patients were more likely to develop lymphopenia (57.1%) than mild patients (34.5%), which is consistent with previous studies that severe patients have higher rates of lymphopenia^[6,10,13,18]^. Lymphopenia is also observed in SARS-CoV and MERS-CoV infected patients but with different mechanisms^[19]^.

Consistent with previous study^[20]^, we also observed the decreased absolute number of CD3^+^ T cells in severe and mild patients (85.7%vs51.7%); moreover, the number of both CD4^+^ and CD8^+^ T cells were markedly lower in severe patients than mild patients. So far, the potential mechanisms of lymphopenia, particularly the decrease of CD4^+^ and CD8^+^ T cell counts in COVID-19 patients remain unclear, the SARS-CoV-2 might have a direct influence on lymphocytes or lymphatic organs, or through an indirect pathway^[12,13]^. The corresponding mechanisms warrant further investigation.

Previous studies of COVID-19 patients have demonstrated that cytokine storm syndrome (CRS) could happen in SARS-CoV-2 infected patients^[9,10]^. Cytokine storms, which can quickly induce organ failure and threaten the life of the patients, are regarded as an important cause of death in COVID-19 patients. Recent studies have reported an increase in serum cytokine levels in COVID-19 patients, especially in severe patients, and suggest that cytokine storm is associated with disease severity^[6,13]^. In our study, more severe patients occurred with the elevated IL6 (64.3%) and IL10 (64.3%) levels than mild patients (IL6: 34.5%; IL10: 13.8%).

Furthermore, the levels of IL6 and IL10 in severe group were significantly higher than mild group. Further analysis demonstrated that the variations of IL6 were closely related to the disease progression of COVID-19 patients. This result is consistent with a previous study, which suggests IL6 as an effective target for disease monitoring and treatment ^[14,21]^. IL6, as an acute phase inflammatory cytokine, plays an important role in regulating inflammatory responses. Elevated IL6 level is often observed in the patients with acute respiratory distress syndrome^[22]^and may be related with T lymphocyte damage. However, the underlying mechanisms of IL6 and other cytokines involved in COVID-19 patients needs further investigation.

Our results indicated that, as an infection-related factor, ESR was also significantly higher in severe patients compared to mild patients, which is consistent with previous studies^[9,23]^. ESR is a simple laboratory test for evaluating the inflammatory or acute response state. Previous studies have reported that symptoms and signs plus ESR can predict pneumonia effectively^[24]^. ESR also can be used as a monitoring factor in diagnosis and management of Gemcitabine-induced pulmonary toxicity^[25]^. Furthermore, ESR is reported as a powerful predictor for coronary heart disease^[26,27]^. However, the exact role of ESR in COVID-19 patients is still controversial, as a study from Gong et al. indicate that ESR is unrelated to the disease severity in patients with COVID-19 pneumonia^[28]^.

Early examination of risk factors in COVID-19 patients could distinguish severe patients from mild patients effectively and facilitate appropriate treatment. Recent studies have reported several factors that may be related to the severity of COVID-19 patients, including IL6^[14]^, lymphopenia^[12]^, NLR^[17]^ and N8R^[13]^. Although we also observed a higher frequency of occurrence of lymphopenia in severe patients, it was not a potential factor for predicting the severity of COVID-19 patients according to LASSO binary logistic regression analysis. Liu et al. Have indicated that N8R has even better performance than NLR in the ROC curve analysis^[13]^. In our study, N8R was also significantly higher in severe patients, unfortunately, we cut it out since there are no results in LASSO binary logistic regression analysis. Our results demonstrated that the potential markers to identify severe patients were ESR, D-D, age, ALB and IL6 by LASSO binary logistic regression analysis and ESR had the best performance with a higher AUC value in the ROC curve analysis. Therefore, ESR can be used as a powerful indicator to predict disease severity in SARS-CoV-2 infected patients.

In conclusion, this study provides a correlation between clinical parameters and disease severity of COVID-19 patients. Our study of mild and severe patients demonstrates that these clinical parameters, including ESR, D-D, age, ALB and IL6 are closely related with the disease progression of COVID-19 patients. Among these parameters, ESR has the best performance and can be used as an effective indicator to predict the severity of disease. Our study may help to identify severe COVID-19 patients in early stage, in order to provide supportive treatment and reduce the mortality.

## Data Availability

All data and note links are available from the authors

## Acknowledgments

This work is supported by the Introduction of Talent Program of Chongqing Medical and Pharmaceutical College (ygz2019304) and Joint Projects of Chongqing Science and Technology Commission and Health Commission (2019MSXM023).

